# Interim estimates of increased transmissibility, growth rate, and reproduction number of the Covid-19 B.1.617.2 variant of concern in the United Kingdom

**DOI:** 10.1101/2021.06.03.21258293

**Authors:** John Dagpunar

## Abstract

This paper relates to data from the Wellcome Sanger Institute, UK, regarding Covid-19 genomic surveillance. We use a simple model to give point estimates of the effective reproduction numbers of the B.1.617.2 and B.1.1.7 lineages in England, from sequenced data as at 15 May 2021. Comparison with the estimated reproduction number of B.1.1.7 enables an estimate of the increased transmissibility of B.1.617.2. We conclude that it is almost certain that there is increased transmissibility that will rapidly lead to B.1.617.2 becoming the prevailing variant in the UK. The derived estimates of increased transmissibility have uncertainty relating to the actual distribution of the generation interval, but they do point, under present conditions of vaccination coverage and NPIs, to exponential growth of positive cases.

## 1. Sequencing data

Table 1 shows numbers and proportions of positive genomically sequenced cases in England that are B.1.617.2 (Indian variant) and B.1.1.7 (Kent variant), for the period 20 March to 22 May 2021. That data [1], is expressed in cases per week (by date on which sample is collected), calculated as two-week averages centred on 27 March, 3 April, …., 8 May, 15 May. It excludes recent travellers, and special studies, so that it more realistically reflects community transmission. Until mid-April 2021 it excluded “surge testing” but this is now included as it is no longer thought to bias the proportion of the now common B.1.617.2 variant. The aim of this paper is to obtain a rough estimate of the effective reproduction numbers of these two variants at 15 May 2021, and hence the increased transmissibility of B.1.617.2 over B.1.1.7, for different distributions of the generation interval. The data shown in the first 6 columns of table 1, extracted from [1], relate to these two lineages which account for between 98% and 99% of all cases in this period. In tandem with these data, we used the number of all positive cases in England (sequenced and un-sequenced), by date of specimen [2], and averaged over a centred 15-day cycle to replicate the approach used for sequenced data.

**Table 1:**
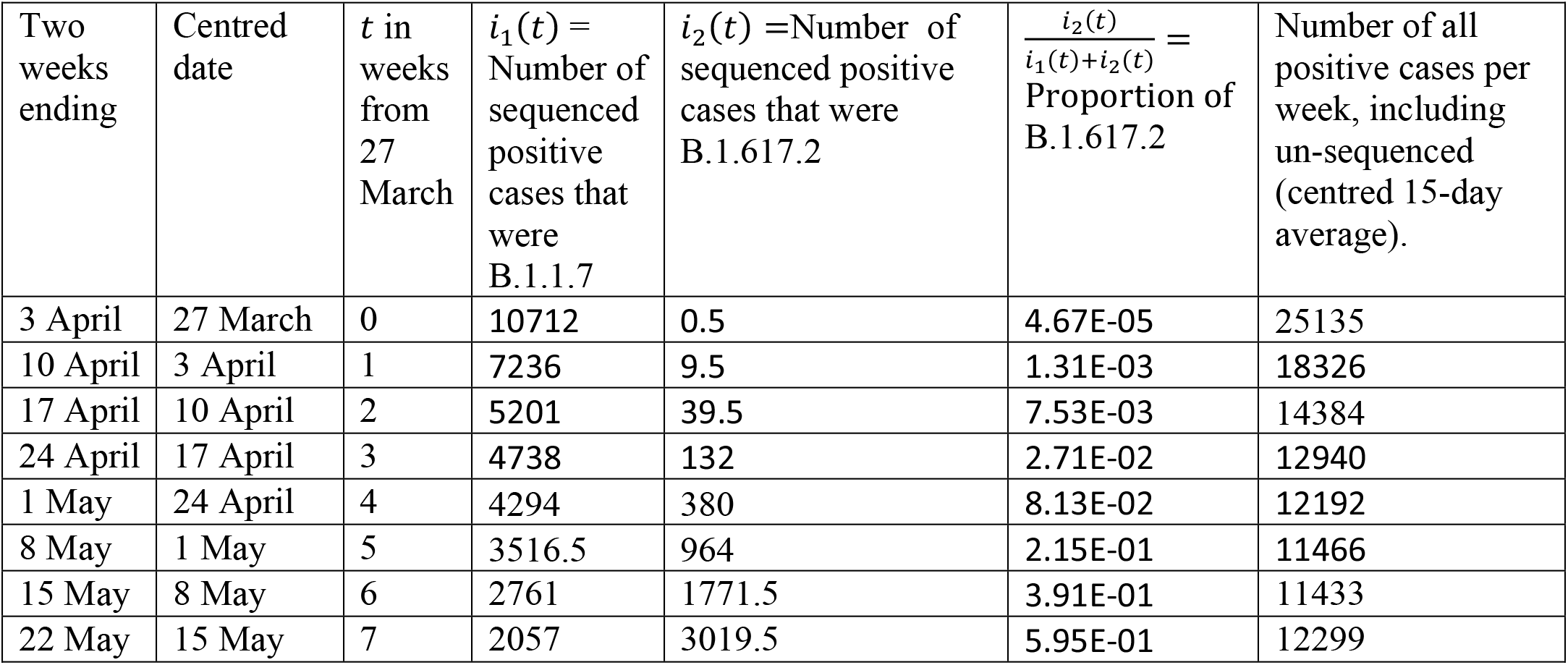
Sequencing data for B.1.1.7 and B.1.617.2.

| Two weeks ending | Centred date | $t$ in weeks from 27 March | $i_1(t)$ = Number of sequenced positive cases that were B.1.1.7 | $i_2(t)$ = Number of sequenced positive cases that were B.1.617.2 | $\frac{i_2(t)}{i_1(t)+i_2(t)}$ = Proportion of B.1.617.2 | Number of all positive cases per week, including un-sequenced (centred 15-day average). |
| --- | --- | --- | --- | --- | --- | --- |
| 3 April | 27 March | 0 | 10712 | 0.5 | 4.67E-05 | 25135 |
| 10 April | 3 April | 1 | 7236 | 9.5 | 1.31E-03 | 18326 |
| 17 April | 10 April | 2 | 5201 | 39.5 | 7.53E-03 | 14384 |
| 24 April | 17 April | 3 | 4738 | 132 | 2.71E-02 | 12940 |
| 1 May | 24 April | 4 | 4294 | 380 | 8.13E-02 | 12192 |
| 8 May | 1 May | 5 | 3516.5 | 964 | 2.15E-01 | 11466 |
| 15 May | 8 May | 6 | 2761 | 1771.5 | 3.91E-01 | 11433 |
| 22 May | 15 May | 7 | 2057 | 3019.5 | 5.95E-01 | 12299 |

Relevant work in this area is that of Public Health England [3]. That includes estimates of secondary attack rates for the two variants and of the growth rate of B.1.617.2 relative to that of B.1.1.7 at 22 May 2021. The date is relevant as the present paper shows that the growth rate of B.1.617.2 appears to be decreasing. Similar questions were addressed regarding the detection in South East England in September 2020 of the, then, more transmissible variant B.1.1.7., [4].

## 2. Effective reproduction numbers, comparative transmissibility, and growth rate

Let *R*_1_(*t*) and *R*_2_(*t*) denote the effective reproduction numbers of B.1.1.7 and B.1.617.2 at time *t*. Based upon those cases that were sequenced, the sample proportion of all cases at time *t*, sequenced and un-sequenced, that are variant *j*, is 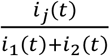. Here, it is assumed that those cases that are sequenced are representative of the larger population of all positive cases. With the Sanger data, the exclusion of travellers and special studies makes this more likely. The estimated incidence of variant *j* cases per day is therefore 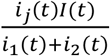. where *I*(*t*) is the 15-day centred average incidence of all cases, sequenced and un-sequenced, on day *t*. By conditioning on the incidence at *t* - *u*, we have to a good approximation,

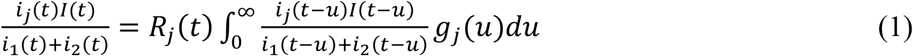

where *g*_*j*_ (*u*) is the probability density function of the generation interval for variant *j*. Therefore,

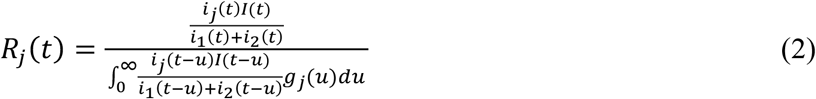

for *j* = 1,2. It is well known that case incidence will experience exponential growth with rate *λ*_*j*_ (*t*) say, and so the last equation can be rewritten as

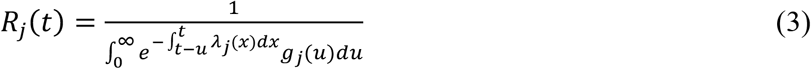

Which may be compared with

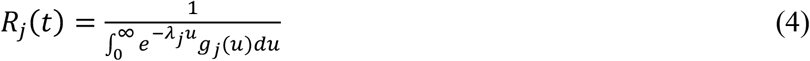

as in [5], for the case of a constant growth rate *λ*_)_.

A generation interval is the time between a (primary) individual becoming infected and infecting (secondary) individuals. I assume that the distribution of generation interval is stationary and is the same for both B.1.1.7 and B.1.617.2. The latter will be the case providing the transmission rate of one variant is simply a scaled version of the other. Assuming this to be so, the increase in transmissibility of B.1.617.2 over B.1.1.7 is 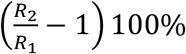. I assume that the generation interval is gamma distributed, as for example in [6,7], with mean *μ*, shape parameter *α*, and standard deviation 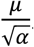. For *α* >1, this allows for an initial phase following infection in which there is a monotonic increase in transmission rate approximating to the latent and pre-symptomatic period, a second phase of high transmission, followed by a third one of monotonic decrease.

## 3. Results

The Sanger data is available as centred two-week averages every week. This was converted to daily data by linear interpolation within each week, of the proportions of cases that are variant 1 and 2. The daily number of all positive cases was obtained from [2], calculating centred 15-day averages, for each of the 49 days (7 weeks). Table 2 shows point estimates, as of 15 May 2021, of *R*_1_(49) and *R*_2_(49), the increased transmissibility, and the overall reproduction number

**Table 2.** Reproduction numbers and increased transmissibility as of 15 May 2021.

| (Mean, standard deviation) of generation interval in days | $R_1(49)$ | $R_2(49)$ | $R(49)$ | Increased transmissibility |
| --- | --- | --- | --- | --- |
| (5.2,1.72) | 0.750 | 1.37 | 1.12 | 83% |
| (3.78,0.91) | 0.789 | 1.23 | 1.05 | 56% |
| (6.78,3.93) | 0.703 | 1.51 | 1.19 | 115% |
| (3.95,1.51) | 0.788 | 1.25 | 1.06 | 59% |
| (3.01,0.74) | 0.820 | 1.17 | 1.03 | 43% |
| (4.91,2.97) | 0.761 | 1.34 | 1.10 | 76% |

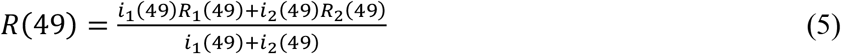

The selected parameter values for specimen gamma distributed generation intervals are those given in [7] for studies on infected individuals in Singapore and Tianjin, China.

In table 3, the sample proportion of variants obtained from sequenced data is used to construct weekly estimates of variant case incidence in the population. From this table, the weekly growth rates are calculated. For example, the growth rate for B.1.617.2 in week 5 is ln 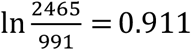. From this table the doubling or halving times may be calculated. For example, the doubling time of B.1.617.2 at 15 May 2021 (centred date) is 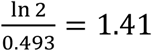 weeks. The dynamically changing growth rates are shown in figure 1

**Table 3.**
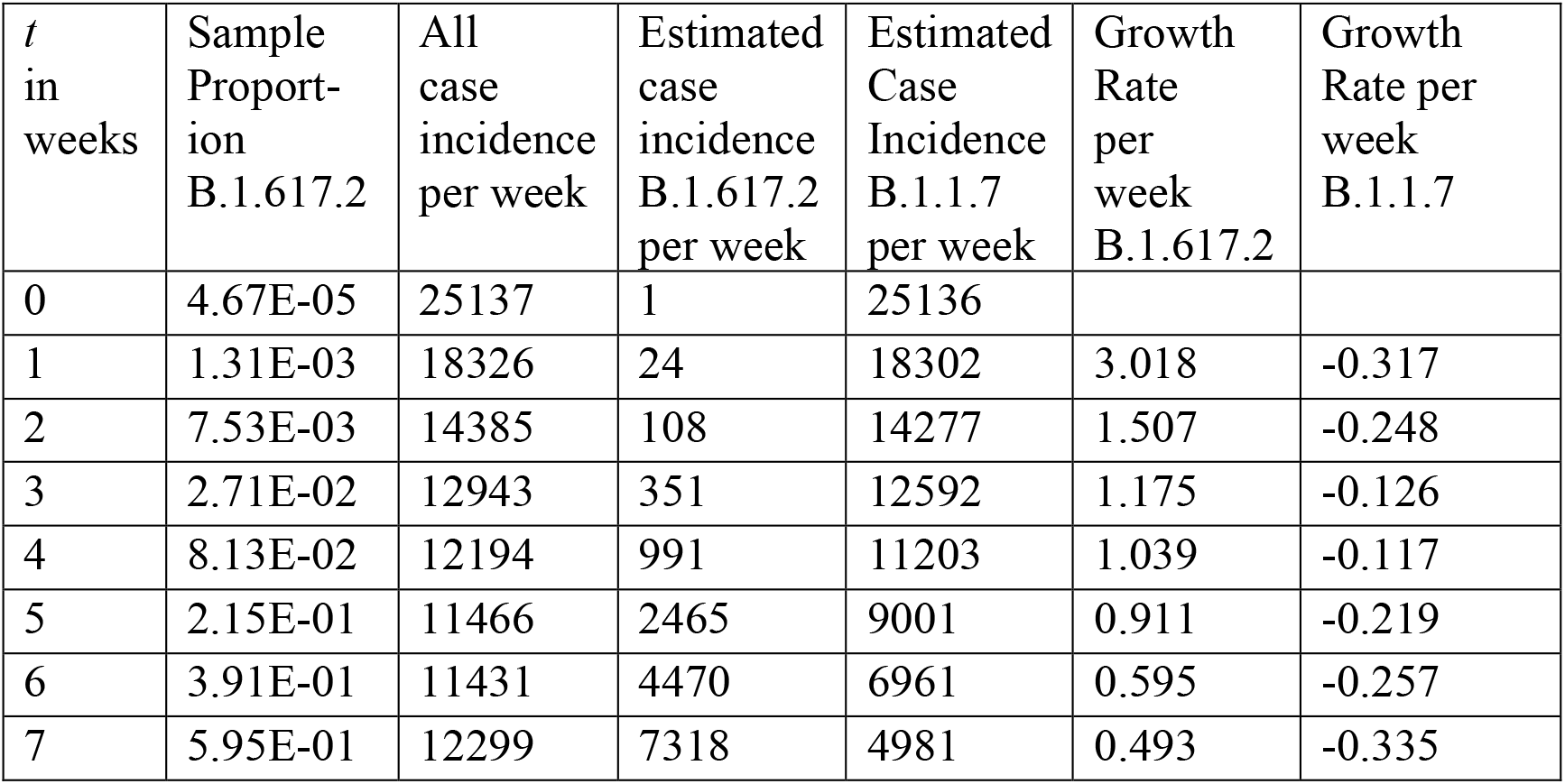
Weekly estimates of variant case incidence and growth rates.

**Figure 1:**
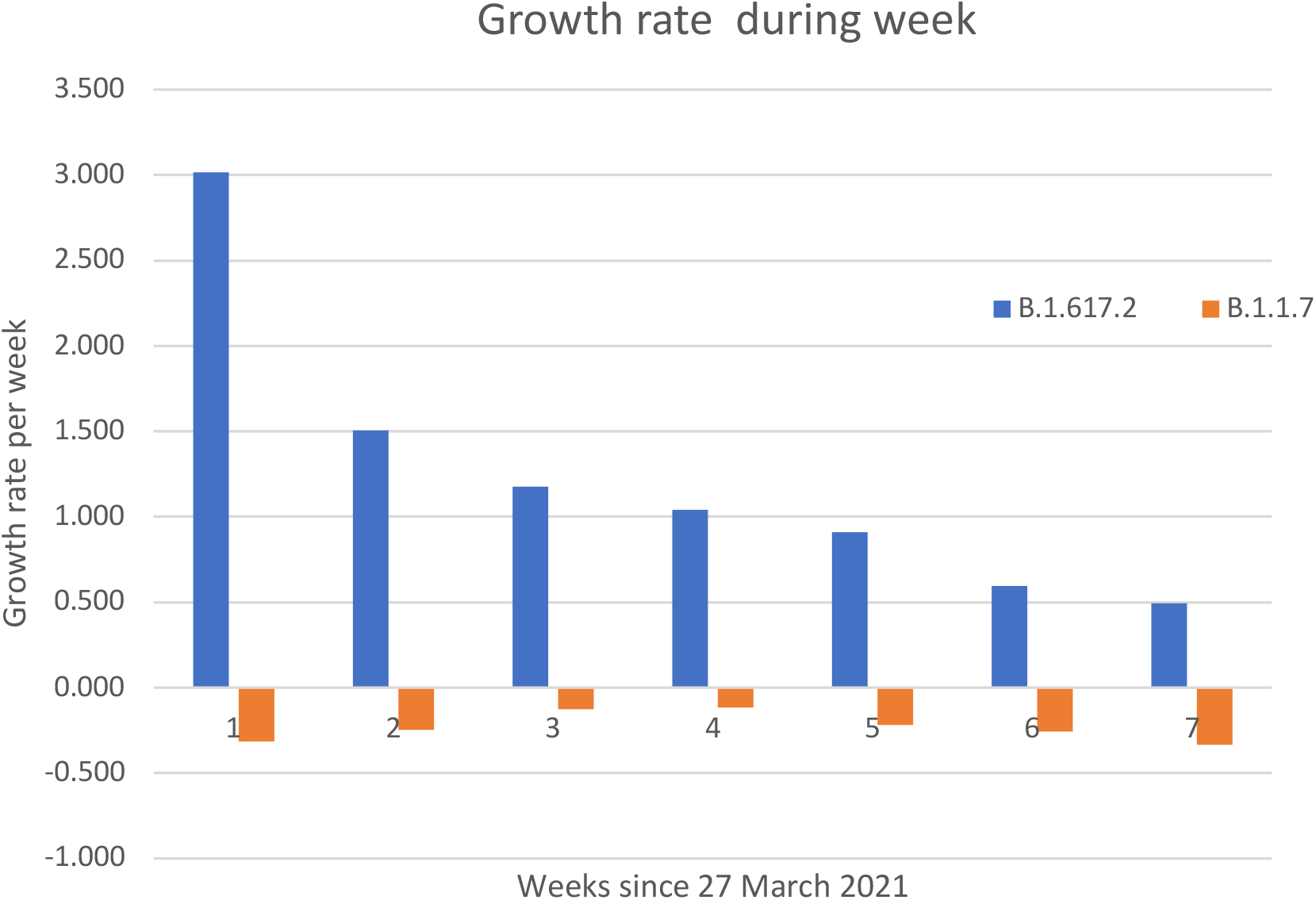
The Growth rate per week since 27 March 2021

## 4. Discussion and Conclusions

The degree of increased transmissibility is large and varies considerably with the assumed distribution of generation interval. Although not shown here, for a given mean generation interval, transmissibility increase was relatively insensitive to variability in generation interval. These results might be compared with secondary attack rates among contacts that have not travelled [3], (13.5% for B.1.617.2 versus 8.1% for B.1.1.7). Most of the results in the final column of table 2 are larger than those suggested by the SAGE update of 13 May 2021, [8], where a 50% increase in transmissibility was thought to be a realistic possibility. For comparison, the official overall effective reproduction number [9] was mainly in the range (0.8-1.1) during this period.

The estimates of increased transmissibility of B.1.617.2 depend upon the distribution of the generation interval and *t*. We have shown only the values calculated for all the data up to *t* = 7, that is 15 May (centred date). For low values of *t* there is great uncertainty in the growth rate of B.1.617.2 (but not of B.1.1.7) because the incidence is so low. However, it was found that by *t* = 6, 7 the incidence was now high enough for the increased transmissibility to have stabilised and so the results shown in the last column of table 2 are a reasonable guide for each given distribution of generation interval.

These results are contingent upon the validity of the model assumptions including the assumption that the sequencing data are representative of overall community transmission. With regards to this, local outbreaks might first become apparent in environments where the transmission rate is biased to high values, for example in communities with large average household size and less than average social distancing. When B1.617.2 becomes the almost exclusive variant, as seems inevitable, then its proportion of all cases becomes 1, which removes uncertainty about the extent to which sequenced data is representative of the entire population. The uncertainty about the distribution of generation interval remains.

Figure 1 reveals an interesting feature in that the estimated positive growth rate of B1.617.2 has been decreasing and the positive decline rate of B.1.1.7 increasing. A possible explanation for this is that in both cases the increasing vaccine roll-out in the UK is more than compensating for the gradual relaxation of NPIs. Nevertheless, when B.1.617.2 eventually accounts for almost all cases, column 3 of table 2 shows that under present circumstances there will be exponential growth as the reproduction number is significantly above 1. It might be possible to reduce this below 1 by rapidly increasing vaccination coverage, but this might not be enough in a context of further relaxation of NPIs. Policymakers will need to decide whether the risks of not taking anticipatory action should that prove to be necessary, outweigh the risks of intervening should that be shown not to be required. Finally, an unknown is whether increased case numbers will lead to increased hospitalisations and deaths.

## Data Availability

The data used for the analysis are in the public domain, as listed [1] and [2] in the references and with links shown here:
https://covid19.sanger.ac.uk/lineages/raw?show=B.1.617.2%2CB.1.1.7&colorBy=p&lineage=B.1.617.2&area=overview&latitude=50.161569&longitude=-2.591175&zoom=4.33&date=2021-05-22
https://coronavirus.data.gov.uk/details/cases?areaType=nation&areaName=England

